# Cross-cultural adaptation and validation of the Japanese Charité Alarm Fatigue Questionnaire (CAFQa) among ICU nurses and physicians: a multicenter study

**DOI:** 10.64898/2026.04.07.26350292

**Authors:** Tomoo Sato, Mitsuko Ishiseki, Yuki Kataoka, Hidehiro Someko, Hiroki Sato, Kota Minami, Takao Kaneko, Hisashi Takeda, Adam Crosby

## Abstract

**Objectives:** Alarm fatigue is a patient safety concern in ICUs, yet no validated instrument exists to assess alarm fatigue among healthcare professionals in non-Western settings. This study aimed to cross-culturally adapt the Charité Alarm Fatigue Questionnaire (CAFQa) into Japanese and evaluate its reliability and validity among ICU nurses and physicians.

**Methods:** The Japanese CAFQa was cross-culturally adapted following the COnsensus-based Standards for the selection of health Measurement INstruments (COSMIN) guidelines, including forward translation, back-translation, expert panel review, and cognitive interviews. A multicenter cross-sectional validation study was performed across eight ICUs at five hospitals in Japan. A total of 129 participants (103 nurses and 26 physicians) completed the Japanese CAFQa, the NIOSH Brief Job Stress Questionnaire, and the Insomnia Severity Index (ISI). Structural validity, internal consistency, test–retest reliability (n = 102), convergent validity, and known-groups validity were assessed.

**Results:** CFA confirmed the two-factor structure with acceptable fit (CFI = 0.922, RMSEA = 0.041, SRMR = 0.076), with standardized factor loadings ranging from 0.33 to 0.82. The two factors were not correlated (r = 0.05). Cronbach’s alpha was 0.688 for the overall scale, 0.805 for Alarm Stress, and 0.649 for Alarm Coping. Test–retest ICCs ranged from 0.616 to 0.753. The CAFQa total score correlated with the NIOSH total (r = 0.261) and the ISI total (r = 0.338). Healthcare professionals with ≥4 years of ICU experience had higher Alarm Coping scores than those with 1–3 years (median 7.0 vs 6.5), and physicians scored higher on Alarm Coping than nurses (median 8.0 vs 7.0).

**Conclusions:** The Japanese CAFQa demonstrated acceptable structural validity, reliability, and convergent and known-groups validity, providing the first validated tool for quantitatively measuring alarm fatigue in Japan.

**Implications for Clinical Practice:** The Japanese CAFQa enables ICU managers to quantify alarm fatigue at individual and unit levels, identify high-risk staff, and evaluate the effectiveness of alarm management interventions.

## Introduction

The ICU relies extensively on monitoring devices that generate alarms to alert healthcare professionals to changes in patients’ conditions. Of these alarms, 72% to 99% of these alarms are false or do not require clinical intervention [1–4]. This excessive volume of nonactionable alarms leads to alarm fatigue, which has dual consequences. At the patient level, frequent false alarms condition healthcare professionals to disregard alerts, resulting in missed critical alarms and, in some cases, patient deaths [5]. At the staff level, chronic alarm exposure causes headaches, hearing impairment [6], sleep disturbance [7], and is significantly associated with burnout [8]. Alarm fatigue has been recognized as a major patient safety concern, designated as a National Patient Safety Goal in the United States [1] and frequently ranking among the top 10 technology hazards identified by the Emergency Care Research Institute [9].

Reliable measurement instruments that survey both nurses and physicians are essential for addressing this issue. Wunderlich et al. developed the Charité Alarm Fatigue Questionnaire (CAFQa), a 9-item self-report instrument designed for both nurses and physicians [10]. The CAFQa measures alarm fatigue across two dimensions: alarm stress (5 items), assessing the psychosomatic effects of excessive alarms, and alarm coping (4 items), evaluating coping mechanisms and organizational support. Each item is scored on a 5-point Likert scale (0–4).

The cross-cultural applicability of the CAFQa varies across language versions. The construct validity of the original German version was confirmed through confirmatory factor analysis [10], and its two-factor structure was subsequently replicated in a larger German multicenter study [11]. The CAFQa has since been translated and validated in English [12] and Dutch [13]. However, psychometric properties have not been uniform across these studies; internal consistency of the Alarm Coping subscale has been consistently low (α = 0.46–0.68) [10–13], and model fit indices have not uniformly met the most stringent criteria. These findings highlight the need to examine the validity and reliability of the CAFQa across diverse linguistic and cultural contexts. To date, no version of the CAFQa has been translated and validated for use among healthcare professionals in a non-Western setting. Adapted the CAFQa into Japanese would not only enable the quantitative measurement of alarm fatigue in Japanese ICUs but also provide the first evidence of the instrument’s applicability beyond Western European and English-speaking populations.

Therefore, the aim of this study was to translate the CAFQa into Japanese following established cross-cultural adaptation guidelines and to evaluate its reliability and validity among ICU nurses and physicians, including structural validity, internal consistency, test–retest reliability, measurement error, convergent validity, and known-groups validity.

## Methods

### Study design and participants

We followed the methodological framework recommended by COnsensus-based Standards for the selection of health Measurement Instruments (COSMIN) [14]. This was a cross-sectional, multicenter validation study. ICU nurses and physicians working in eight adult ICUs at five hospitals in Japan were eligible for participation. Inclusion criteria were: (1) working as a nurse or physician in an ICU, (2) having at least one month of ICU experience, and (3) ability to read and understand Japanese. Participants with prior exposure to the Japanese CAFQa were excluded.

### Ethical Approval

This study was approved by the institutional review boards of Kobe City College of Nursing (approval number: 25104-04) and Kyoto University Graduate School and Faculty of Medicine (approval number: R5182-1). All participants provided informed consent.

### Translation Process

Two researchers (TS and MI) independently translated the original English CAFQa into Japanese. Discrepancies were resolved through discussion; unresolved disagreements were arbitrated by a third researcher (YK). A native English speaker (CA) with fluency in Japanese, blinded to the original instrument, performed a back-translation. A multidisciplinary expert panel comprising four ICU physicians, four ICU nurses, and one translator evaluated semantic, idiomatic, experiential, and conceptual equivalence, resolving remaining discrepancies through consensus.

### Cognitive interviews

To evaluate face validity, cognitive interviews were conducted using the preliminary Japanese version of the CAFQa. The purpose of cognitive testing was to determine whether the items in the preliminary version were easily understood by the target population. A total of seven participants meeting the eligibility criteria—two ICU physicians and five ICU nurses—were recruited for the cognitive interviews. Participants first completed the preliminary Japanese CAFQa. Individual interviews were then conducted by the researchers to assess the content, ease of responding, and comprehensibility of each item. All participants reported that the items were easily understood and could be answered without ambiguity. No substantial modifications were required, and the final Japanese version of the CAFQa was established [15].

### Data Collection

Data were collected from ICU physicians and nurses across five hospitals (eight ICUs) in Japan. The target sample size was determined based on the COSMIN recommendation that structural validity studies require a minimum of 100 participants [14]. To allow for incomplete responses and dropout during the test–retest phase, we aimed to recruit at least 120 participants. Demographic data including age, sex, professional role, years of clinical experience, years of ICU experience, and highest educational attainment were collected. Participants completed the Japanese CAFQa, the National Institute for Occupational Safety and Health (NIOSH) Brief Job Stress Questionnaire [16], and the Insomnia Severity Index [17]. The NIOSH Brief Job Stress Questionnaire is a validated self-report instrument that assesses multiple dimensions of occupational stress, including quantitative workload, job control, and interpersonal conflict, and was selected to examine the relationship between alarm fatigue and general occupational stress. The ISI is a 7-item self-report measure that evaluates the severity of insomnia symptoms and was included based on prior evidence linking alarm exposure to sleep disturbance in ICU staff. Approximately two weeks later, participants were asked to complete the CAFQa a second time to assess test–retest reliability. Responses were collected via Microsoft Forms.

### Statistical Analysis

Descriptive statistics were used for the demographics and clinical characteristics of the study population. Reliability was evaluated through internal consistency, test–retest reliability, and measurement error. Internal consistency was assessed using Cronbach’s alpha, with a value of 0.7 or greater considered acceptable [18]. Test–retest reliability was examined by calculating the intraclass correlation coefficient (ICC) between the first and second administrations of the Japanese CAFQa, with an ICC of 0.6 or above indicating good reliability [19]. Measurement error was evaluated by calculating the standard error of measurement (SEM) using Bland-Altman plot analysis and the ICC formula [20,21]. The SEM reflects within-subject variability across repeated measurements and is useful for evaluating individual-level change. The minimally detectable change (MDC), which represents the variability in scores attributable to measurement error, was calculated using the following formula: 1.96 × √2 × SEM. The MDC indicates the minimum change required to be considered significant based on the reliable change index [22]. Construct validity was evaluated using confirmatory factor analysis to verify the two-factor structure of the CAFQa: alarm stress and alarm coping. Given the ordinal nature of the 5-point Likert scale items, the ULS estimator based on polychoric correlations was employed, consistent with prior validation studies [10–13]. Model fit was assessed using the CFI, TLI, RMSEA, and SRMR [23]. Convergent validity was assessed by examining correlations between the Japanese CAFQa scores and the NIOSH Brief Job Stress Questionnaire and Insomnia Severity Index scores using Pearson’s correlation coefficient [24], with absolute values below 0.3 interpreted as weak, 0.3 to 0.5 as moderate, and above 0.5 as strong [25]. Known-groups validity was examined using the Mann-Whitney U test. We hypothesized that participants with longer ICU experience would show higher Alarm Coping scores, reflecting the development of alarm management competence over time, and that physicians and nurses might differ in coping-related scores due to differences in decision-making authority regarding alarm settings. CAFQa total scores were not expected to differ between occupations, given the shared alarm environment in ICU settings. Face validity was assessed through the cognitive interviews described above. Statistical significance was set at p < 0.05. All analyses were performed using R version 4.4.3, Psych [26], Lavaan [27], and irr [28] software programs.

### Missing data handling

Participants who did not complete all nine CAFQa items were planned to be excluded from the primary analyses (complete-case analysis). For the convergent validity analyses, participants with missing data on the NIOSH Brief Job Stress Questionnaire or the ISI were excluded pairwise. No imputation methods were applied.

## Results

### Participants

A total of 396 ICU nurses and physicians across the eight ICUs were invited to participate. Of these, 129 completed the initial survey (response rate: 32.6%). The mean (SD) age was 33.1(9.6) years, with 93 (72.1%) being female. The mean occupation experience was 10.2 (8.7) years, and the mean ICU experience was 6.8 (6.5) years. Regarding educational background, 66 (51.2%) held a university degree, 44 (34.1%) had vocational or junior college education, 11 (8.5%) held doctoral degrees, and 8 (6.2%) held master’s degrees (Table 1). For the test–retest reliability analysis, the 129 initial respondents were asked to complete the CAFQa a second time approximately two weeks later; 102 responded (retention rate: 79.1%; overall response rate from the original sample: 26.0% [102/396]). All 129 participants completed all items of the CAFQa, the NIOSH Brief Job Stress Questionnaire, and the ISI at Time 1, with no missing data across any measure.

**Table 1.**
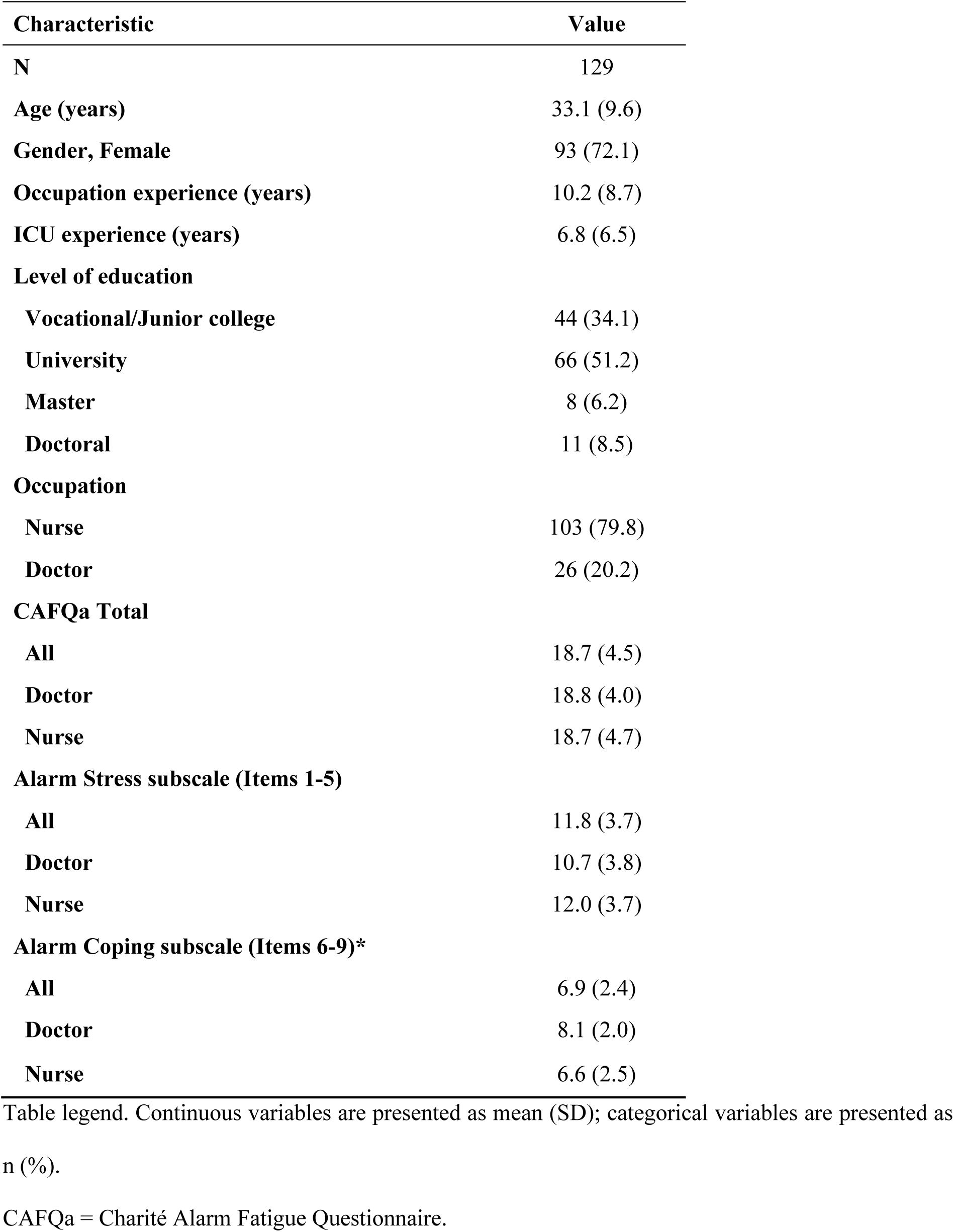
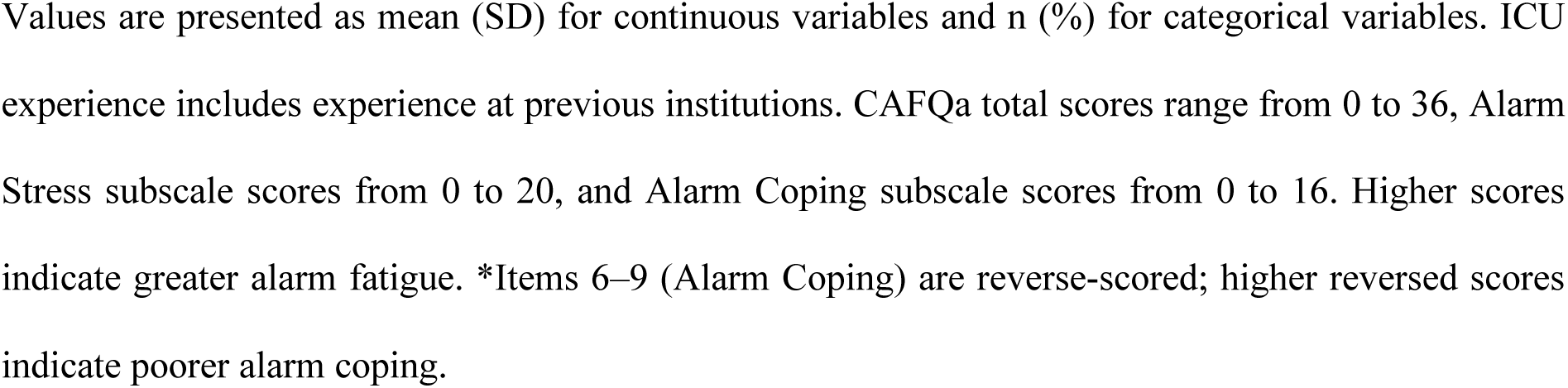
Demographic Characteristics of Participants.

| **Characteristic** | **Value** |
| --- | --- |
| **N** | 129 |
| **Age (years)** | 33.1 (9.6) |
| **Gender, Female** | 93 (72.1) |
| **Occupation experience (years)** | 10.2 (8.7) |
| **ICU experience (years)** | 6.8 (6.5) |
| **Level of education** |  |
| **Vocational/Junior college** | 44 (34.1) |
| **University** | 66 (51.2) |
| **Master** | 8 (6.2) |
| **Doctoral** | 11 (8.5) |
| **Occupation** |  |
| **Nurse** | 103 (79.8) |
| **Doctor** | 26 (20.2) |
| **CAFQa Total** |  |
| **All** | 18.7 (4.5) |
| **Doctor** | 18.8 (4.0) |
| **Nurse** | 18.7 (4.7) |
| **Alarm Stress subscale (Items 1-5)** |  |
| **All** | 11.8 (3.7) |
| **Doctor** | 10.7 (3.8) |
| **Nurse** | 12.0 (3.7) |
| **Alarm Coping subscale (Items 6-9)\*** |  |
| **All** | 6.9 (2.4) |
| **Doctor** | 8.1 (2.0) |
| **Nurse** | 6.6 (2.5) |

Table legend. Continuous variables are presented as mean (SD); categorical variables are presented as n (%).
CAFQa = Charité Alarm Fatigue Questionnaire.
Values are presented as mean (SD) for continuous variables and n (%) for categorical variables. ICU experience includes experience at previous institutions. CAFQa total scores range from 0 to 36, Alarm Stress subscale scores from 0 to 20, and Alarm Coping subscale scores from 0 to 16. Higher scores indicate greater alarm fatigue. \*Items 6–9 (Alarm Coping) are reverse-scored; higher reversed scores indicate poorer alarm coping.

### Reliability

The Cronbach’s alpha coefficient for the overall CAFQa scale was 0.688. The alpha coefficient for the alarm stress subscale was 0.805, and that for the alarm coping subscale was 0.649. Test-retest reliability was assessed in 102 participants. The ICC for the total CAFQa score was 0.709 (95% CI: 0.597–0.794). The ICC for the alarm stress subscale was 0.753 (95% CI: 0.654–0.826), and the ICC for the alarm coping subscale was 0.616 (95% CI: 0.480–0.724). The standard error of measurement (SEM) and the minimal detectable change (MDC) are presented in Table 2. For the CAFQa total score, the SEM was 2.52 and the MDC was 6.98. For the alarm stress subscale, the SEM was 1.91 and the MDC was 5.30. For the alarm coping subscale, the SEM was 1.55 and the MDC was 4.29. The Bland-Altman plots (Figure 1) showed that the majority of data points fell within the limits of agreement (±1.96 SD) for all three scores. Item-level descriptive statistics are presented in Supplementary Table S1. Potential floor or ceiling effects, defined as more than 15% of respondents selecting the lowest or highest possible score, were observed in three items. Item 9 exhibited a floor effect (29.5%), Item 6 exhibited a ceiling effect (22.5%), and Item 4 exhibited a ceiling effect (20.9%). For the reverse-scored Alarm Coping items, floor effects indicate strong coping behaviors, whereas ceiling effects indicate poor coping.

**Figure 1.**
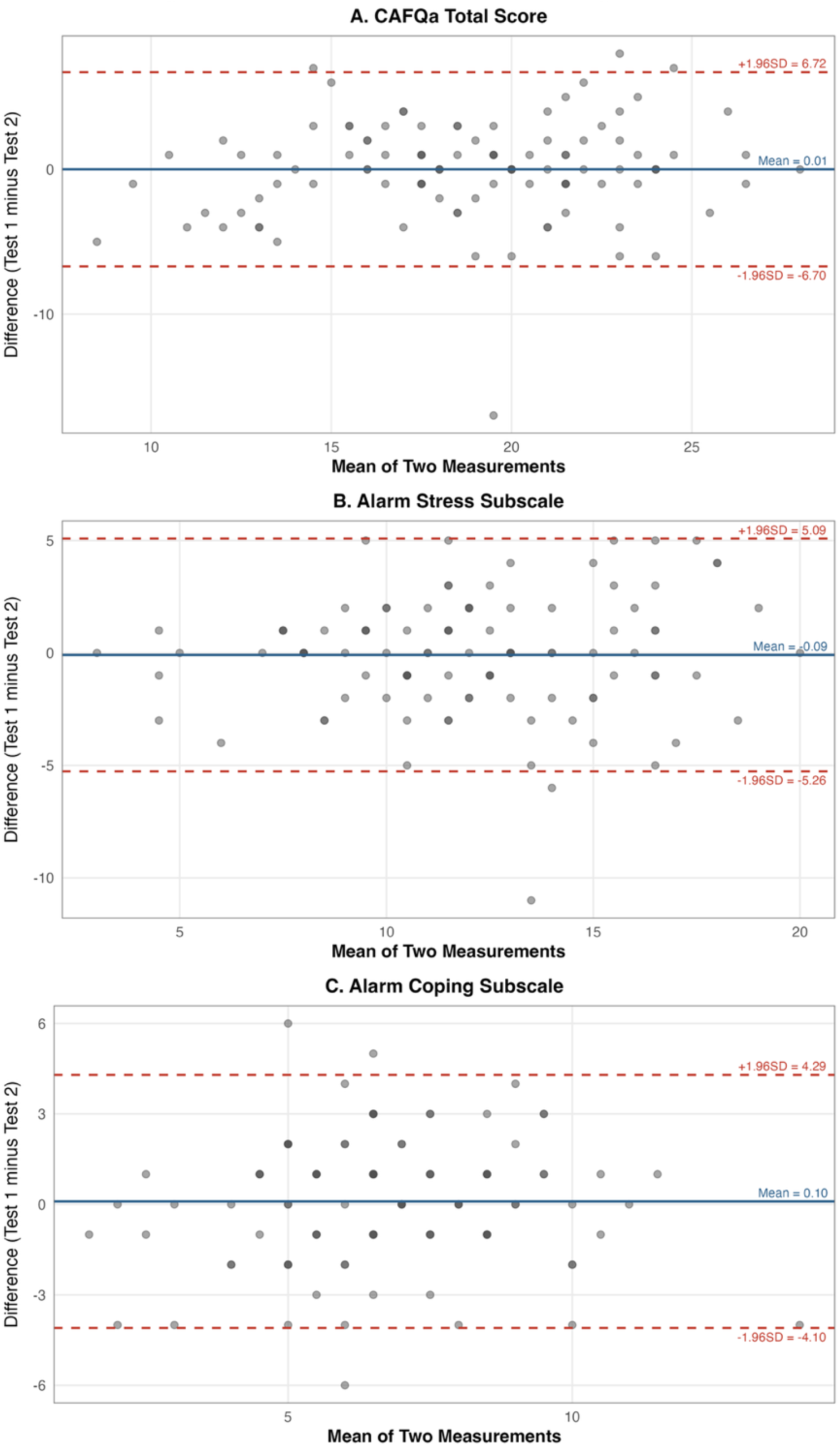
Bland-Altman Plots for Test–Retest Agreement of the Japanese CAFQa (n = 102) Figure legend. Panel A shows the CAFQa total score, Panel B shows the Alarm Stress subscale, and Panel C shows the Alarm Coping subscale. The x-axis represents the mean of the two measurements (Test 1 and Test 2), and the y-axis represents the difference between the two measurements (Test 1 minus Test 2). The solid red line indicates the mean difference, and the dashed red lines indicate the upper and lower limits of agreement (mean ± 1.96 SD). The second assessment was completed approximately two weeks after the initial administration. Alarm Coping subscale scores reflect reverse-scored items (Items 6–9).

**Table 2.** Reliability of the Japanese CAFQa: Internal Consistency, Test–Retest Reliability, and Measurement Error.

| Scale | N_items | Cronbach's<br>alpha | ICC | SEM | MDC |
| --- | --- | --- | --- | --- | --- |
| **CAFQa Total** | 9 | 0.688 | 0.709 (0.597–0.794) | 2.52 | 6.98 |
| **Alarm Stress** | 5 | 0.805 | 0.753 (0.654–0.826) | 1.91 | 5.30 |
| **Alarm Coping** | 4 | 0.649 | 0.616 (0.480–0.724) | 1.55 | 4.29 |

ICC = intraclass correlation coefficient; SEM = standard error of measurement; MDC = minimal detectable change at the 95% confidence level, calculated as $1.96 \times \sqrt{2} \times \text{SEM}$ .
Cronbach's alpha $\geq 0.70$ was considered acceptable for internal consistency. ICC $\geq 0.60$ was considered indicative of acceptable test–retest reliability. Test–retest reliability was assessed in 102 participants who completed the CAFQa approximately two weeks after the initial administration.

### Validity

The two-factor model of the Japanese CAFQa was evaluated using CFA with the ULS estimator.based on polychoric correlations. The model fit indices were: CFI = 0.922; TLI = 0.891; RMSEA = 0.041; SRMR = 0.076 (Supplementary Table S2). For the alarm stress factor, loadings ranged from 0.55 to 0.80, and for the alarm coping factor, from 0.33 to 0.82 (Figure 2). The correlation between the factors was weak (r = 0.05), indicating that the two factors were independent.

**Figure 2.**
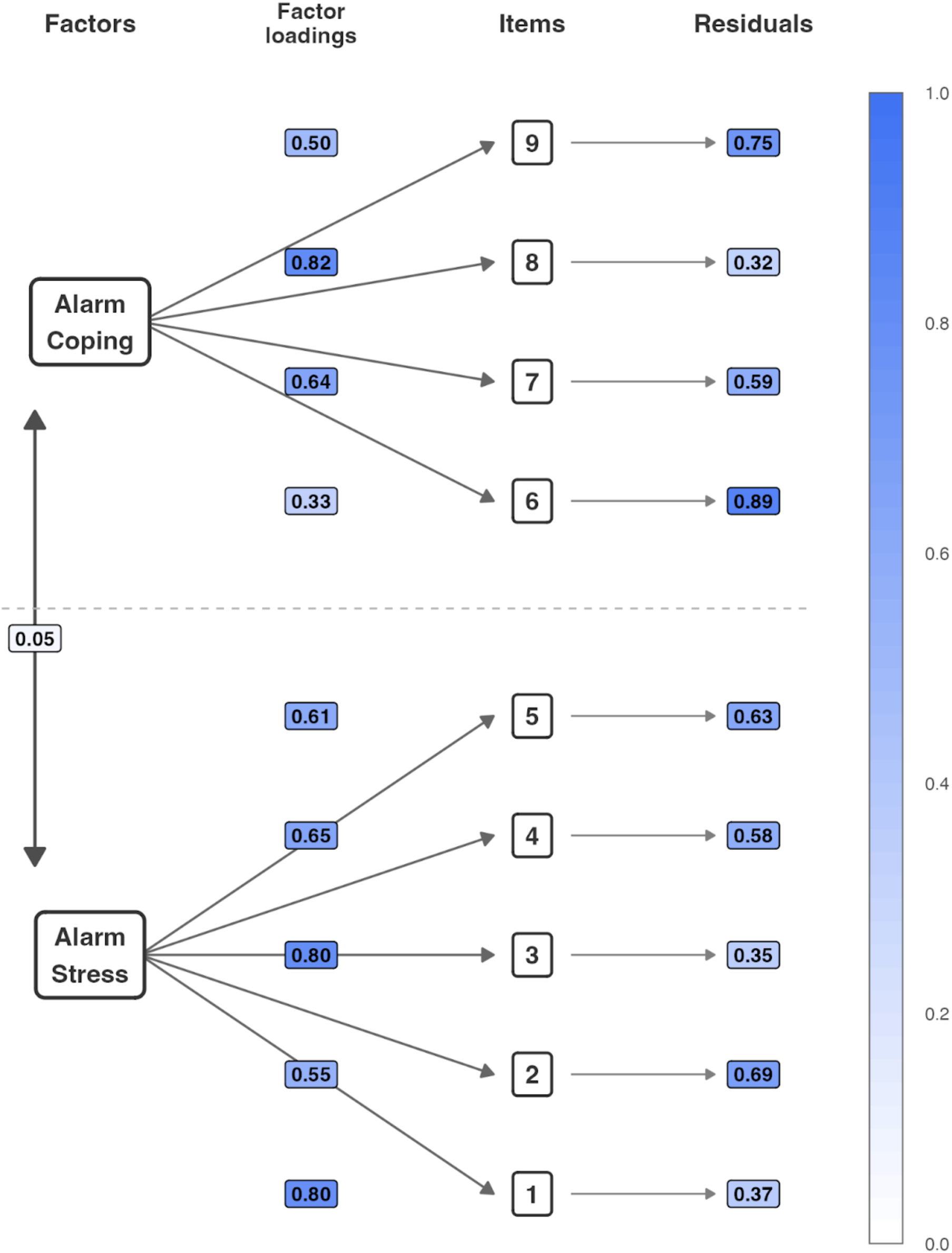
Structural diagram of the two factor model. Figure legend. The diagram displays the two-factor model comprising Alarm Stress (Items 1–5) and Alarm Coping (Items 6–9). Values on the left represent standardized factor loadings, and values on the right represent residual variances for each item. The inter-factor correlation between Stress and Coping was 0.05. The color gradient on the right side of the figure represents the magnitude of residual variance, with darker shading indicating higher values. The model was estimated using the MLR estimator. All factor loadings were statistically significant (p < 0.001).

The CAFQa total score correlated with the NIOSH total score (r = 0.261, p = 0.003) and the ISI total score (r = 0.338, p < 0.001) (Figure 3). The alarm stress subscale correlated with the ISI total (r = 0.369, p < 0.001) and the NIOSH total (r = 0.256, p = 0.004). The alarm coping subscale was not correlated with the NIOSH total (r = 0.094, p = 0.288) or the ISI total (r = 0.063, p = 0.476). The CAFQa total score correlated with the NIOSH quantitative workload subscale (r = 0.189, p = 0.032). The Alarm Coping subscale was selectively correlated with workplace relational and environmental factors (interpersonal conflict, physical environment, supervisor and coworker support; r = 0.218–0.233, p < 0.01–0.05), but not with quantitative workload, job control, or skill utilization (Supplementary Table S3). The Alarm Coping subscale also correlated with ICU experience (r = 0.267, p = 0.002).

**Figure 3.**
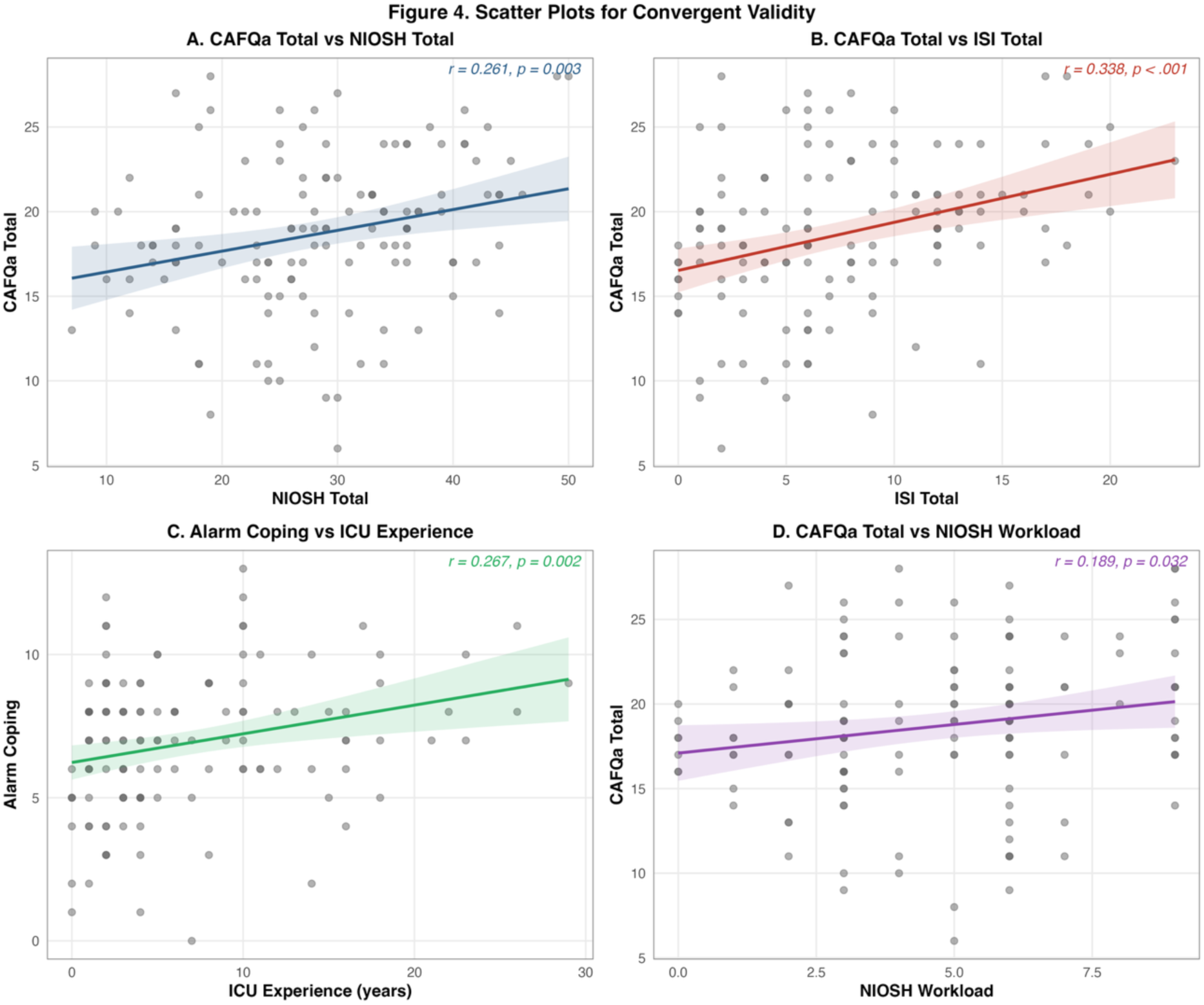
Scatter Plots for Convergent and Hypothesis-Specific Validity of the Japanese CAFQa. Figure legend. CAFQa = Charité Alarm Fatigue Questionnaire; NIOSH = National Institute for Occupational Safety and Health Brief Job Stress Questionnaire; ISI = Insomnia Severity Index. Panel A shows the correlation between the CAFQa total score and the NIOSH Brief Job Stress Questionnaire total score. Panel B shows the correlation between the CAFQa total score and the Insomnia Severity Index (ISI) total score. Panel C shows the correlation between the Alarm Coping subscale score and years of ICU experience. Panel D shows the correlation between the CAFQa total score and the NIOSH quantitative workload subscale score. Each data point represents an individual participant. Solid lines indicate fitted regression lines, and shaded areas represent 95% confidence intervals. Pearson’s correlation coefficients (r) and p values are displayed in each panel. Alarm Coping subscale scores reflect reverse-scored items (Items 6–9), with higher scores indicating poorer alarm coping.

For known-groups validity, healthcare professionals with ≥4 years of ICU experience had higher CAFQa total scores (p = 0.039) and Alarm Coping subscale scores (p = 0.044) than those with 1–3 years of experience (Supplementary Table S5). A similar pattern was observed for occupation experience, with those having ≥4 years showing higher CAFQa total scores (p = 0.007) and Alarm Coping scores (p = 0.003). The CAFQa total score did not differ between physicians and nurses (p = 0.986), whereas physicians scored higher on the Alarm Coping subscale than nurses (p = 0.003).

## Discussion

This study translated the CAFQa into Japanese following established cross-cultural adaptation guidelines and evaluated the reliability and validity of the Japanese CAFQa among ICU nurses and physicians across eight ICUs at five hospitals in Japan. To our knowledge, this is the first validated instrument for quantitatively measuring alarm fatigue among healthcare professionals in Japan. Confirmatory factor analysis supported the hypothesized two-factor structure, and the instrument demonstrated acceptable reliability. Furthermore, the a priori hypotheses regarding known-groups validity were largely supported: healthcare professionals with longer ICU experience showed higher Alarm Coping scores, and physicians scored higher on Alarm Coping than nurses, consistent with expected differences in alarm management competence and decision-making authority. These findings provide evidence that the Japanese CAFQa is a valid and reliable tool for quantifying alarm fatigue to inform alarm management strategies in Japanese ICU settings.

### Structural and Consistency Validity

Confirmatory factor analysis supported the hypothesized two-factor structure comprising Alarm Stress and Alarm Coping, with acceptable model fit (CFI = 0.922; RMSEA = 0.041; SRMR = 0.076) and all standardized factor loadings reaching statistical significance. These fit indices are consistent with the English validation study (CFI = 0.957; RMSEA = 0.079; SRMR = 0.077) [12] and comparable to prior German and Dutch studies [10,11,13]. These findings are consistent with prior validation studies. Factor loadings for the Alarm Stress subscale ranged from 0.55 to 0.80, all exceeding the recommended threshold of 0.50 [10–13]. However, Item 6, which addresses the regular updating and sharing of alarm response protocols, demonstrated a low factor loading of 0.33 with a high residual variance of 0.80, indicating that only approximately 12% of the variance in this item was explained by the Coping factor. Prior studies [10–13] have reported factor loadings for Item 6 ranging from 0.21 to 0.53, suggesting that this is an inherent structural limitation of the original scale rather than a translation-specific issue. While the other Coping items primarily assess personal or unit-level alarm management capabilities, Item 6 captures organizational process management, a qualitatively distinct construct that may account for its persistently low factor loading. The CFI TLI, and SRMR fell short of the most stringent COSMIN criteria (CFI ≥ 0.95; TLI ≥ 0.95; SRMR < 0.06) [14], though the degree of deviation was relatively modest and likely attributable in part to the low factor loading of Item 6. Prior studies [10–13] have also reported model fit indices that did not uniformly satisfy the most stringent criteria, suggesting that the values observed here are within the range of the existing literature.

A notable finding was the high factor loading of Item 8 (0.82), which assesses whether alarms clearly identify the patient, room, and urgency level. This was substantially higher than the loadings of 0.35– 0.45 reported in prior studies [10,11,13]. Notably, the English version also demonstrated a relatively high factor loading for Item 8 (0.74) [12], suggesting that the importance of alarm information clarity in coping perception may not be unique to the Japanese context but rather a feature that becomes more prominent outside the original German-speaking setting. These findings suggest that the ability to rapidly distinguish clinical intervention alarms from non-actionable ones constitutes the core of perceived coping capacity. This finding points to the potential importance of alarm system information clarity as a target for future coping-related interventions.

The inter-factor correlation between Stress and Coping was r = 0.05, indicating virtual independence. A similar pattern was observed in the English validation study (r = 0.12) [12], whereas moderate correlations were reported in the German (r = 0.40–0.60) [10,11] and Dutch (r = 0.46) [13] versions. The low inter-factor correlations observed in the Japanese and English versions suggest that the independence of the two subscales may not be unique to the Japanese context. When alarm information clarity is high, clinicians may acknowledge the stress of frequent false alarms while maintaining confidence in their ability to identify important alarms, effectively decoupling stress from coping perception. This independence suggests that intervention targeting stress reduction and coping enhance may need to be considered. Cultural and linguistic factors may also have contributed to this finding, and formal measurement invariance testing is warranted.

### Reliability

The Alarm Stress subscale demonstrated good internal consistency (α = 0.805), comparable to the original study. The Alarm Coping subscale yielded a lower alpha of 0.649, below the conventional threshold of 0.70 [14]. This may reflect the small number of items (k = 4), which inherently constrains internal consistency estimates. Notably, this value exceeded those reported in the original German studies (α = 0.46–0.57) [10,11,13] and was comparable to the English version (α = 0.68) [12], despite the persistent weakness of Item 6.

Test–retest reliability was adequate, with ICCs of 0.709 for the total score, 0.753 for Alarm Stress, and 0.616 for Alarm Coping. The Bland-Altman analysis confirmed that the majority of data points fell within the limits of agreement. The MDC values—6.98 for the total score, 5.30 for Alarm Stress, and 4.29 for Alarm Coping—provide reference values for future intervention studies to distinguish genuine changes from measurement error. However, given the near-zero inter-factor correlation (r = 0.05), the interpretation of the CAFQa total score as a unidimensional summary of alarm fatigue should be made with caution, as it may obscure distinct patterns of alarm stress and alarm coping. The total score may serve as a useful screening indicator, but subscale-level interpretation is recommended for clinical and research purposes.

### Convergent and Known-Groups Validity

The Japanese CAFQa demonstrated weak to moderate correlations with the NIOSH Brief Job Stress Questionnaire (r = 0.261) and the Insomnia Severity Index (r = 0.338). Although modest, these values are consistent with the specificity of alarm fatigue as a construct that overlaps with, but is not synonymous with, general occupational stress or sleep disturbance. The Alarm Stress subscale showed stronger correlations with both measures than the Alarm Coping subscale, further supporting the construct distinction between the two factors.

Known-groups comparisons provided additional validity evidence. Healthcare professionals with greater ICU experience had higher Alarm Coping subscale scores, which may indicate greater competence in alarm management developed over time, although survivor or retention effects—whereby professionals with inherently higher coping capacity are more likely to remain in the ICU—cannot be excluded in this cross-sectional design. Physicians scored higher on Alarm Coping than nurses, potentially reflecting differences in decision-making authority regarding alarm settings, while the two groups did not differ on the total score, consistent with the shared alarm environment in ICU settings. Notably, the Alarm Coping subscale was not correlated with occupational stress or sleep disturbance, in contrast to the Alarm Stress subscale. This dissociation, consistent with the near-zero inter-factor correlation (r = 0.058), is also in line with the English version, which reported a similarly low inter-factor correlation (r = 0.12) [12]. These findings suggest that the two subscales capture distinct dimensions: Alarm Coping reflects organizational and procedural aspects of alarm management rather than individual stress responses. This interpretation may be further informed by Alarm Coping’s selective correlations with workplace relational and environmental factors (r = 0.218–0.233) and its differentiation between physicians and nurses (p = 0.003), although the small physician subgroup (n = 26) warrants cautious interpretation. These findings suggest that coping-targeted interventions may need to consider organizational factors as well as individual stress reduction.

### Limitations

This study has some limitations. First, the physician subgroup was small (n = 26), limiting the statistical power of occupational comparisons; the observed differences should be interpreted as preliminary evidence. The internal consistency of the Alarm Coping subscale (α = 0.649) fell below the conventional threshold of 0.70; however, this value was comparable to the English version (α = 0.68) [12] and exceeded the German (α = 0.46–0.57) [10,11] and Dutch (α = 0.46) [13] versions, suggesting an inherent characteristic of the coping construct rather than a translation-specific issue. Second, the TLI (0.891) marginally fell below the recommended threshold of 0.90, although the CFI (0.922), RMSEA (0.041), and SRMR (0.076) were all within acceptable ranges, suggesting that further model refinement may be warranted with larger samples. Third, the convergent validity correlations with NIOSH occupational stress and ISI scores were modest, which may partly reflect the specificity of alarm fatigue as a construct distinct from general occupational stress or sleep disturbance. Fourth, the overall response rate was 32.6%, raising the possibility of non-response bias; nurses and physicians with stronger concerns about alarm fatigue may have been more likely to participate, potentially affecting the generalizability of the findings.

## Conclusion

This study translated, cross-culturally adapted, and validated the Japanese version of the Charité Alarm Fatigue Questionnaire (CAFQa) among ICU nurses and physicians across multiple hospitals. The Japanese CAFQa provides a brief, validated instrument for the objective assessment of alarm fatigue among Japanese ICU healthcare professionals. This tool enables the quantification of alarm fatigue at both individual and institutional levels, facilitating the identification of high-risk groups and the evaluation of alarm management interventions. Future studies should examine the responsiveness of the Japanese CAFQa to alarm reduction programs and its applicability in broader clinical settings.

## Data Availability

The raw data are not publicly available due to ethical and privacy restrictions.

https://doi.org/10.17605/OSF.IO/D46QM

## Clinical trial registration

Not applicable.

## Acknowledgements

We extend our gratitude to Professor Maximilian Markus Wunderlich for granting us permission to translate the original version of the CAFQa. We also acknowledge Mr. Kenjiro To, Mr. Ryo Hirai, Mr.Tomohiko Takahashi, Ms. Tsubasa Mori, Mr. Kazuhiro Miura, Mr. Ryota Imanaka, and Mr. Daichi Fujimoto for their contribution to the cognitive interviews.

## Ethics statement

This study was conducted in accordance with the Declaration of Helsinki. The study protocol was approved by the institutional review board of Kobe City College of Nursing (approval number: 25104-04) and Kyoto University Graduate School and Faculty of Medicine (approval number: R5182-1). Participation was voluntary and anonymous. All participants provided informed consent by checking a consent box on the online survey prior to participation.

## CRediT authorship contribution statement

Tomoo Sato: Conceptualization, Methodology, Validation, Formal analysis, Investigation, Data curation, Writing – original draft, Writing – review & editing, Supervision. Mitsuko Ishiseki: Conceptualization, Methodology, Investigation, Data curation. Yuki Kataoka: Methodology, Validation, Formal analysis, Writing – review & editing, Supervision. Hidehiro Someko: Methodology, Validation, Formal analysis, Writing – review & editing, Supervision. Hiroki Sato: Methodology, Investigation, Writing – review & editing. Kota Minami: Methodology, Investigation, Writing – review & editing. Takao Kaneko: Methodology, Investigation, Writing – review & editing. Adam Crosby: Writing – review & editing. Hisashi Takeda: Investigation, Writing – review & editing.

